# The Pandemic Response Commons

**DOI:** 10.1101/2022.06.20.22276542

**Authors:** Matthew Trunnell, Casey Frankenberger, Bala Hota, Troy Hughes, Plamen Martinov, Urmila Ravichandran, Nirav S. Shah, Robert L. Grossman, The Pandemic Response Commons Consortium

## Abstract

**Objective:** A data commons is a software platform for managing, curating, analyzing, and sharing data with a community. The Pandemic Response Commons is a data commons designed to provide a data platform for researchers studying an epidemic or pandemic.

**Methods:** The pandemic response commons was developed using the open source Gen3 data platform and is based upon consortium, data, and platform agreements developed by the not-for-profit Open Commons Consortium. A formal consortium of Chicagoland area organizations was formed to develop and operate the pandemic response commons.

**Results:** We developed a general pandemic response commons and an instance of it for the Chicagoland region called the Chicagoland COVID-19 Commons. A Gen3 data platform was set up and operated with policies, procedures and controls based upon NIST SP 800-53. A consensus data model for the commons was developed, and a variety of datasets were curated, harmonized and ingested, including statistical summary data about COVID cases, patient level clinical data, and SARS-CoV-2 viral variant data.

**Discussion and conclusion:** Given the various legal and data agreements required to operate a data commons, a pandemic response commons is designed to be in place and operating at a low level prior to the occurrence of an epidemic, with the activities increasing as required during an epidemic. A regional instance of a Pandemic Response Commons is designed to be part of a broader data ecosystem or data mesh consisting of multiple regional commons supporting pandemic response through sharing of regional data.

## Background and Significance

A data commons is a software platform for managing, harmonizing, analyzing, and sharing data with a community (1). We describe the Pandemic Response Commons (PRC), which is an open source platform for rapidly setting up data commons for a consortium of researchers, public health workers and decision makers in response to an epidemic or pandemic. In particular, we describe an instance of the Pandemic Response Commons called the Chicagoland COVID-19 Commons (CCC) that serves the Chicago region, and, more broadly, the state of Illinois and surrounding regions.

The CCC contains clinical data from about 90,000 subjects, over 5,300 viral genomes, and public data from a variety of sources.

There were many efforts to collect COVID related data since the start of the COVID-19 pandemic. These include required data collection as part of local and state public health reporting, required data collection as required by federal statutes and regulations, including by the Center for Disease Control (CDC), research related efforts by various NIH Institutes and Centers (2), volunteer efforts. Important projects that provided COVID-19 related data include projects by the John Hopkins University Center for Systems Science and Engineering, the New York Times, and the COVID Tracking Project. Data was also released by companies, for example, the mobility data collected, processed and released by Google. Today, there are many NIH supported projects that are collecting COVID and Long COVID related data, including the NIH Post-Acute Sequelae of SARS-CoV-2 infection (PASC) Project and the National COVID Cohort Collaborative (N3C) (2).

An important role of a data commons is to curate, integrate and harmonize data for a specific community, generally for research purposes. As can be seen from the COVID Tracking Project (3) and the CDC COVID Tracking Data (4), there are important regional differences in the incidence, fatalities, and health disparities associated with COVID-19. This suggests the utility in collecting, integrating and analyzing COVID-19 related data at the local and regional levels, and then aggregating the results. The Pandemic Response Commons is a data platform designed to support this type of local, regional and federated analysis.

## Methods

### Consortium governance

The Pandemic Response Consortium is a private-public partnership that is operated by the Open Commons Consortium (OCC), part of an independent 501(c)(3) not-for-profit corporation that operates several other data commons, including the BloodPAC Data Commons (5) and the Veterans Precision Oncology Data Commons (6).

The Open Commons Consortium provides templates for Consortium Membership Agreements, Data Contributors Agreements, Data Use Agreements, Intellectual Property Agreements, and related agreements so that 1) a consortium can be established to build and operate a data commons, 2) data can be contributed to a data commons, and 3) members of the consortium can form working groups to analyze data in the commons and develop applications, notebooks and software services to enhance the functionality of the commons.

Currently the Pandemic Response Commons has four working groups: the Clinical Data Working Group, the Epidemiological Modeling Working Group, the Community Engagement and Outreach Working Group, and the Data Commons Working Group.

### Data governance

Consortium members that contributed clinical data first obtained IRB approval. The IRBs submitted by members were based upon a common template developed by the consortium. The Clinical Data Working Group met biweekly for several months to develop and refine the data model that each consortium member contributing data used to provide data.

In addition to clinical data, the PRC collected several other types of data from PRC Members, including aggregate counts for different types COVID related incidences, and sequencing data about viral variants. The PRC also collected public data of interest to the COVID research community, including case and fatality data, mobility data, public clinical and imaging data about COVID cases.

### Clinical data and its harmonization

The Clinical Data Working Group developed a data model with over 60 specific data elements to support data modeling efforts with a particular focus on disparities and comorbidities. Adapted from the CDC case report form for persons under investigation developed early in the pandemic, the requested data elements include demographics, symptoms, comorbidities, and lab results. In addition, basic information about a patient’s encounter was requested, including whether the patient was admitted to the hospital, whether they were transferred to the ICU, and whether they were placed on a ventilator and for how long.

The Clinical Data Working Group had to agree on precise definitions of a COVID patient (based on results from one or more of a specified set of laboratory tests) and COVID encounter, defined as a hospital visit within 30 days of a positive COVID test. The definitions used internally by the healthcare organizations varied significantly across the partners. The group also had to agree on precise lists of clinical data codes (ICD-10 codes) describing comorbidities and laboratory results.

The requested data include race and ethnicity where available. This allows questions of disparities in outcomes and variation in occurrence of symptoms and comorbidities to be studied. These data elements are being reported quarterly by participating healthcare partners for all COVID patients.

### Security and compliance

The PRC follows the policies, procedures and controls for a NIST SP 800-53 release 4 Moderate system (7). It also undergoes a yearly penetration test, and a yearly security and compliance audit by a third party.

## Results

### Consortium membership

Currently, the Chicagoland instance of the Pandemic Response Commons Consortium includes 8 members, each of which has signed consortium membership agreements. The consortium members are: Rush University Medical Center, NorthShore University HealthSystem, University of Chicago, University of Illinois at Chicago, St. Anthony Hospital, Sinai Chicago, CommunityHealth, and Southern Illinois University.

Rush University Medical Center, Northshore University HealthSystem, the University of Chicago, the University of Illinois at Chicago, Sinai Health System, St. Anthony Hospital, and others have contributed clinical data to the Chicagoland instance of the PRC. In total, the PRC contains clinical data from over 90,000 subjects with COVID. All controlled access data was contributed under the PRC Data Contributors Agreement under an IRB or with an IRB exemption and can be shared with other consortium members, and, in aggregated form, with other researchers.

### Working group activities

CCC activities are organized into five working groups, each of which has a charter. These were the Clinical Data Working Group, the Epidemiological Modeling Working Group, the Community Engagement Working Group, the Variant Surveillance Working Group, and the Commons Operations Working Group.

The Clinical Data Working Group developed a common data model for the commons and worked with each member to contribute data in the required format. In addition to clinical data, the CCC also collected aggregated counts for cases, deaths, and selected comorbidities that are used by the Epidemiological Modeling Working Group to understand health disparities and to build predictive models.

The Epidemiological Modeling Working Group initially developed hierarchical Bayesian models that predicted future COVID case and fatality counts by county in Illinois. The models were adapted from the models developed by Imperial College(8). These models were updated weekly. The Epidemiological Modeling Working Group also developed a generative Bayesian for the Chicago region that is currently running daily. The Working Group has also developed regression models to understand race/ethnicity differences in case/fatality ratios based on temporal and age-related factors.

The Variant Surveillance Working Group has collected over 5,300 SARS-CoV-2 and contributed them to the PRC, as well as to other national and international virus variant databases.

### Pandemic Response Commons

The PRC was developed with the open source Gen3 software platform (https://gen3.org/). Once the Data Working Group defines a data model or updates an existing data model, the Gen3 software platform automatically generates a data commons with APIs for data access, metadata access, data submission, and authorization and authentication. In this way, the data commons protects controlled access data and makes both controlled access and open access data findable, accessible, interoperable, and reusable (FAIR) (9).

Currently, there is one regional instance of the Pandemic Response Commons (https://pandemicresponsecommons.org) – the Chicagoland COVID-19 Commons. An instance of the PRC includes a portal for exploring, submitting data, and browsing publicly accessible PRC Jupyter Notebooks. The PRC Notebook Browser includes over a dozen Jupyter notebooks that are built over data from the PRC, including case and fatality data, open access clinical data, mobility data, and COVID-19 imaging data.

### Patient level data

To date, three institutions (NorthShore University HealthSystem, Rush University Medical Center, and University of Chicago Medical Center) have submitted Patient Summary Reports with patient-level data starting from March 1, 2020. The PRC has focused on analyzing submitted information for validation and to identify data quality issues, such as differences in format and missing data. Quality analysis included developing plots that compared patient counts by demographic characteristics, presenting symptoms, events (ventilation, ICU admission, etc.), comorbidities, and complications. The following information should be considered illustrative. In particular, because institutions identify COVID patients based on a rule (evidence of positive test result within 30 days of admission), the counts of patients may be different from counts provided by institutions (which, in general, would use different criteria).

### Summary level data

PRC statistical summary reports (SSR) contain aggregate counts in different categories. As aggregate data, they are easier for contributing organizations to share, and, at a sufficiently high level of aggregation, are open access. Count level data from the SSR and other sources are used in various ways by the PRC, including to develop epidemiological models, to provide information for map overlays that are shared with the public, and to provide information for data that is “returned to the community” as part of the activities of the Community Engagement and Outreach Group.

Three institutions (NorthShore University HealthSystem, Rush University Medical Center, and University of Chicago Medical Center) have submitted some form of Statistical Summary Reports of aggregate data. As with the PSR reports, the PRC is analyzing the SSR reports that it has received, assessing for data quality and consistency. The specification for the SSR report is still evolving, partly in response to findings from data quality analysis. The PRC is working with institutions to finalize the SSR specification, after which institutions would resubmit historical SSR data for all days since March 1, 2020.

### SARS-CoV-2 variant data

The PRC has been working with Southern Illinois University (SIU) on a project to analyze the genomic sequence of SARS-CoV-2 virus isolated from samples from patients that test positive for COVID (10). By studying the distribution of genomic variants over time we can better understand the spread of the disease across the state. In addition, in partnership with Illinois Department of Health (IDPH), we have worked to identify the appearance of specific “Variants of Concern” (VOCs) (11).

The project has sequenced over 5,300 SARS-CoV-2 genomes over the course of the pandemic. The genomes span 16 viral clades and include more than 150 variants with sufficient resolution to illuminate the evolution of the viral population in Illinois (Figure 2).

**Figure 1.**
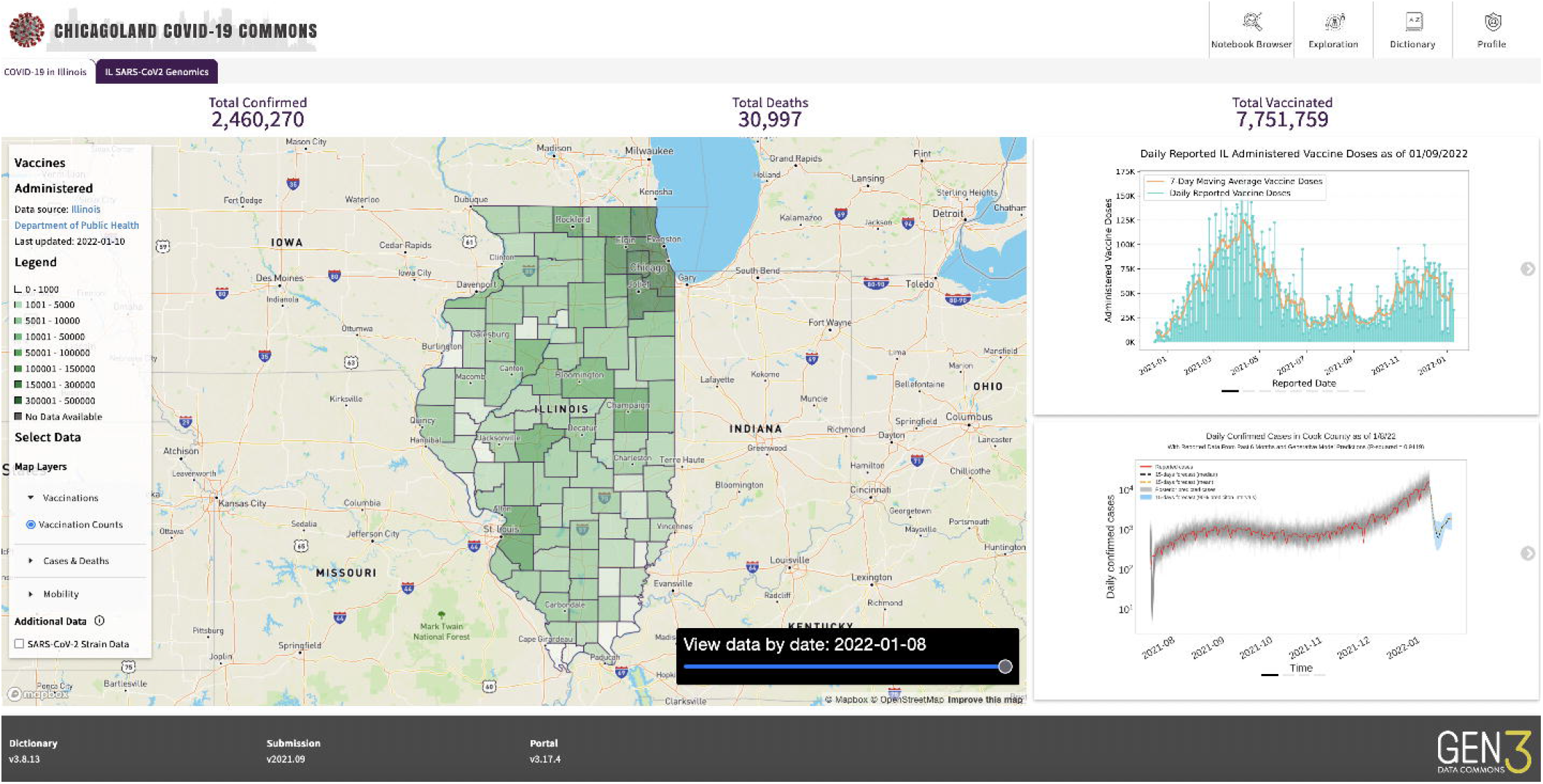
Screenshot of PRC

**Figure 2.**
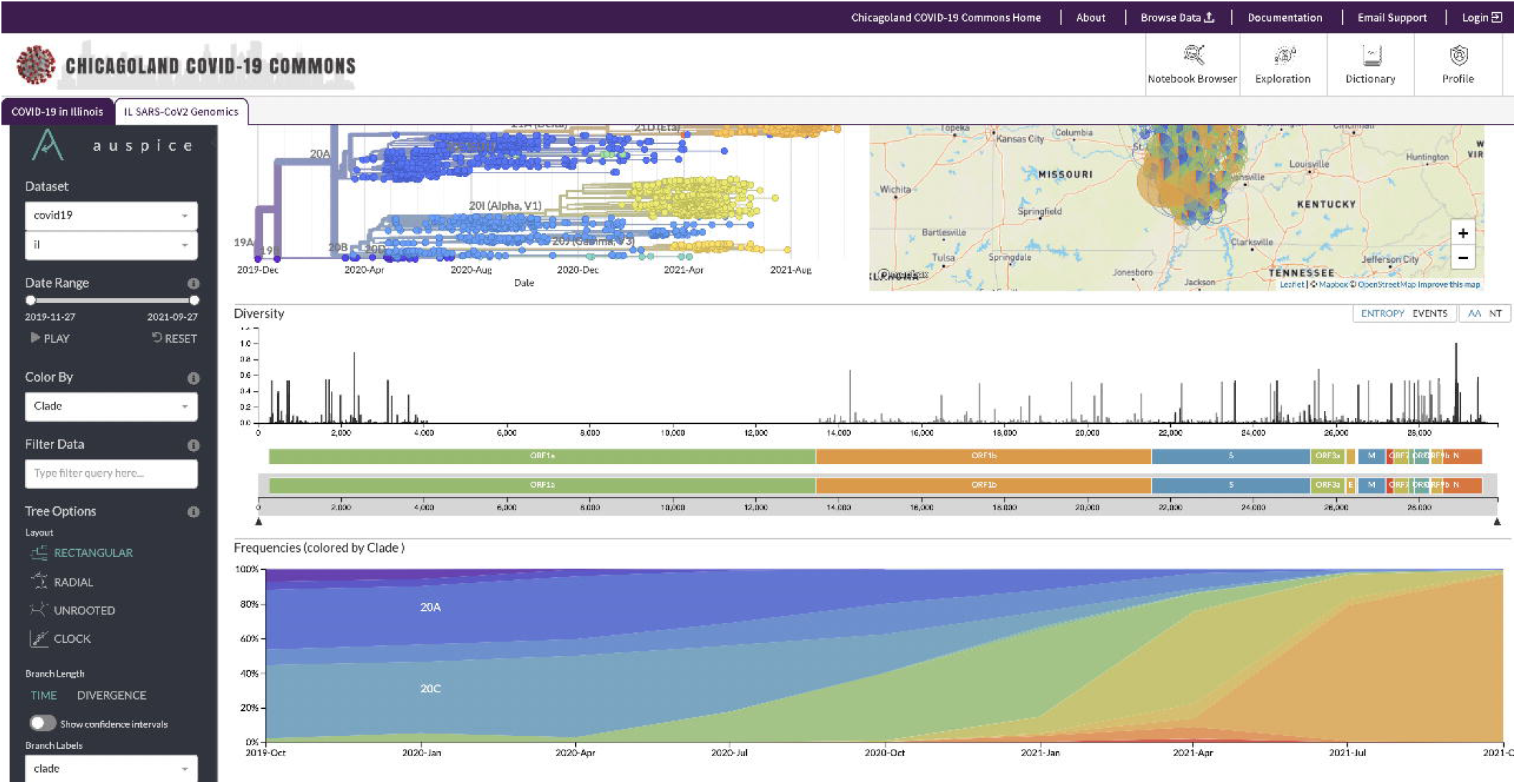
Screenshot of viral variants and their geographic distribution

**Figure 3.**
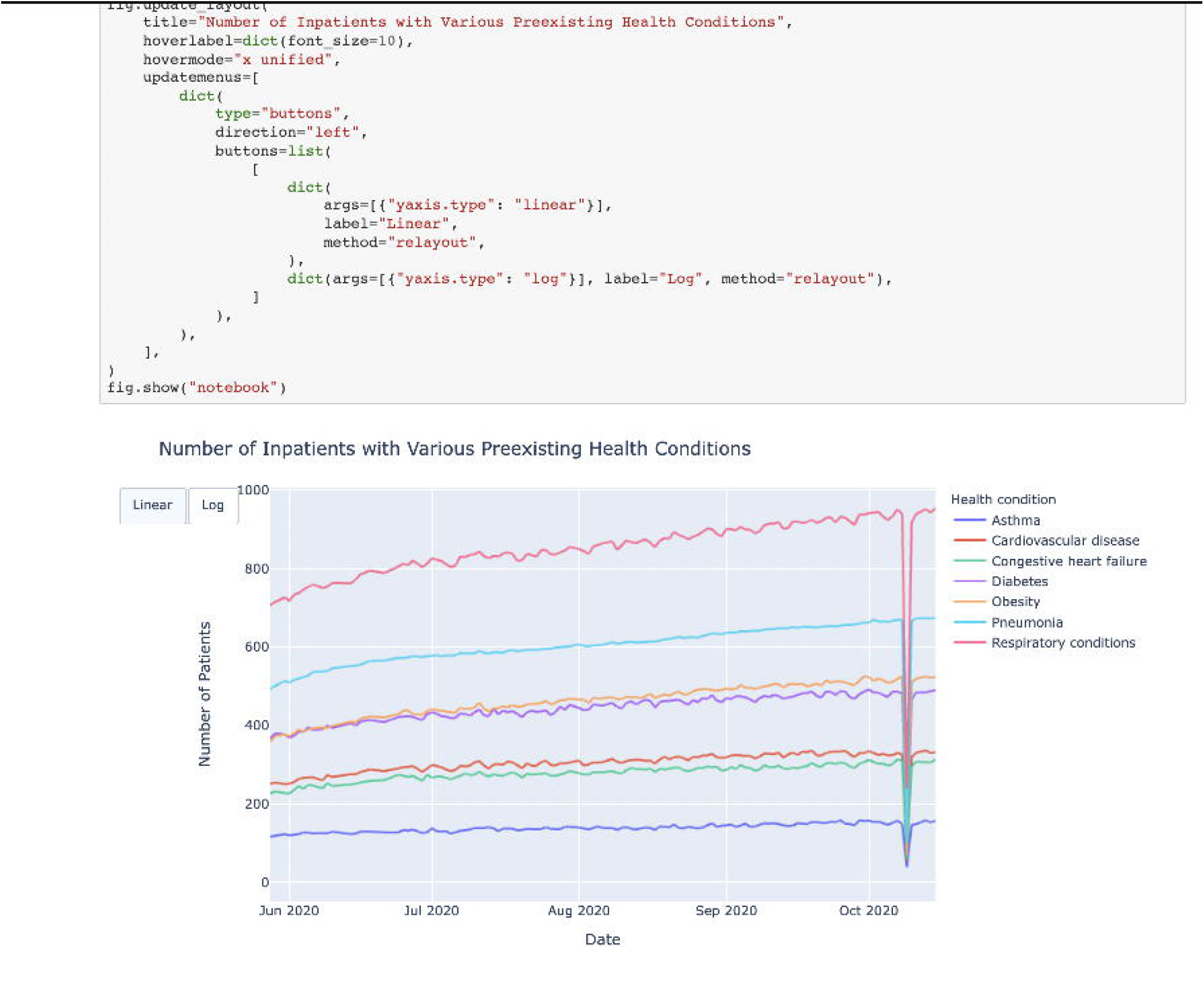
Screenshot involving Jupyter notebook and its analysis.

## Discussion

### Regional COVID commons

There are several important ongoing efforts to gather data at the national level to support clinical research, policy development and decision making at the national scale (2). To drive regional decision making, a regional COVID commons can be an impactful complement to these national efforts for several reasons. First, local data can be richer, since a regional commons can collect, analyze and share some data within its own community that might not be so readily available or of interest to a broader community. Second, regional commons can more easily reflect community concerns about data collection and data use compared to national commons. Third, regional commons can potentially make decisions that make sense for a local community, but may not be as relevant or as practical at a national level.

### Interoperating regional COVID commons

Over the last several years, technology has been developed so different data commons can interoperate and share data in ways that are safe and compliant with relevant regulations and policies. Data that are open and unrestricted can be shared directly; data that are sensitive or otherwise restricted might be shared in aggregate or summary form. In this way, a national COVID-19 data common can be developed that brings together some of the data in regional data COVID-19 data commons, while leaving some of the more sensitive data in place. Even with data remaining in place in regional data commons, this information can still be used through a federated analysis, in which an analysis is distributed over the data in each commons, and the results are returned to provide an integrated picture. In this way, data from multiple regional data commons can be used to enhance the national view of COVID-19 related issues.

### The importance of federated learning

Regional, and, more granularly, organizational data commons, provide a mechanism for the collection and analysis of data by those most knowledgeable about it. In addition, they can contribute towards federated learning across multiple commons operated by different organizations and consortia. The Gen3 data commons allow workflows in dockerized containers to be executed locally, with the results reviewed if necessary, and returned for aggregation.

### Preparatory for the next epidemic and pandemic

A large part of the effort in building the Pandemic Response Commons and the Pandemic Response Commons Consortium was putting in place all the necessary legal agreements (Consortium Membership Agreements, IRBs, Data Contributor Agreements, Data Use Agreements, Intellectual Property Agreements, etc.) so that members could contribute data to the commons. Now that these legal agreements are in place, the commons can be relatively easily used for monitoring for the emergence of new pathogens, and new epidemics and pandemics if they occur in the future. In other words infrastructure such as the PRC can be thought of as part of a preparatory infrastructure to make communities more resilient to future outbreaks.

### Engaging with the community

Another important aspect of the PRC is engaging with the community through the Community Engagement and Outreach Working Group. In the words of one of the PRC members, since data is “from the community” it should be “for the community.”

## Conclusion

The PRC is data commons for the Chicagoland and Illinois region to accelerate research in COVID and Long COVID. It contains clinical data from over 90,000 patients that have experienced COVID, statistical summary reports supporting the analysis of COVID-19 health disparities, over 5,300 COVID-19 viral variants, and a variety of COVID related public data. Data is available to PRC consortium members and in aggregated form to a broader community. Over 12 Jupyter notebooks, a number of map overlays, and several epidemiological models have been developed as part of the project.

The PRC is designed to interoperate with other regional commons containing COVID data as they are developed and to support distributed and federated machine learning of COVID related data.

## Data Availability

The work described in the paper did not generate data nor analyze data but developed and operated a software platform called the Pandemic Response Commons (PRC). Open access data is available through the platform's APIs. Computations over the PRC's controlled access data can be done by submitting Docker containers to the PRC.

https://pandemicresponsecommons.org

## Author Contributions

All the authors contributed to the implementation and/or operations of the Pandemic Response Commons.

## Data Availability Statement

The work described in the paper did not generate data nor analyze data but developed and operated a software platform – the Pandemic Response Commons (PRC). Open access data is available through the PRC’s APIs. Computations over the PRC’s controlled access data can be done by submitting Docker containers to the PRC.

## Figures and Tables

Table 1. Data in the PRC.

**Table 1.** Patient level data in the Chicagoland COVID-19 Commons

| **Institution** | **Number of patients in submission** | **Date of latest submission** |
| --- | --- | --- |
| NorthShore University HealthSystem | 49,181 | February 2022 |
| Rush University Medical Center | 21,567 | October 2021 |
| University of Chicago Medical Center | 22,622 | February 2022 |

## Pandemic Response Consortium

### Commons Operations Working Group

Casey A. Frankenberger, PhD, Rush University Medical Center

Robert L. Grossman, PhD, University of Chicago

Bala Hota, MD, Rush University Medical Center

Troy Hughes, BA, University of Chicago

Gina R. Kuffel, BS, University of Chicago

Plamen Martinov, MBA, Pandemic Response Commons

Pauline Ribeyre, MS, University of Chicago

Lea Savatore, MPH, Pandemic Response Commons

Nirav Shah, MD, NorthShore University Health System

Eric S. Swirsky, JD, University of Illinois at Chicago

Matthew Trunnell, MSc, Pandemic Response Commons

### Clinical Data Working Group

Jacob Krive, PhD, University of Illinois at Chicago, Tim Holper, MS, University of Chicago

Pamela T. Roesch, MPH, Sinai Health System

Nirav Shah, MD, NorthShore University Health System

J. Alan Simmons, BS, Rush University Medical Center

Eric Swirsky, JD, MA, University of Illinois at Chicago

### Epidemiological Modeling Working Group

L. Philip Schumm, MA, University of Chicago

Kenneth J. Locey, PhD, Rush University Medical Center

Robert L. Grossman, PhD, University of Chicago

Zhenyu Zhang, PhD, University of Chicago

Mihai Giurcanu, PhD, University of Chicago

### Community Engagement Working Group

Suzet McKinney, DrPH, Sterling Bay

Stephanie D. Willding, MPA, CommunityHealth

Kim Jay, BA, Sinai Health System

Pamela T. Roesch, MPH, Sinai Health System

Eric Swirsky, JD, MA, University of Illinois at Chicago

Lea Salvatore, MPH, Pandemic Response Commons

Robert L. Grossman, PhD, University of Chicago

Michelle B. Hoffman, PhD, P33

### Variant Surveillance Working Group

Keith T. Gagnon, PhD, Southern Illinois University

Koushik Sinha, PhD, Southern Illinois University

Matthew Trunnell, MSc, Pandemic Response Commons

